# A comprehensive evaluation of an artificial intelligence based digital pathology to monitor large-scale deworming programs against soil-transmitted helminths: a study protocol

**DOI:** 10.1101/2023.09.28.23296266

**Authors:** Peter K. Ward, Sara Roose, Mio Ayana, Lindsay A. Broadfield, Peter Dahlberg, Narcis Kabatereine, Adama Kazienga, Zeleke Mekonnen, Betty Nabatte, Lieven Stuyver, Fiona Vande Velde, Sofie Van Hoecke, Bruno Levecke

## Abstract

**Background:** Manual screening of a Kato-Katz (KK) thick stool smear remains the current standard to monitor the impact of large-scale deworming programs against soil-transmitted helminths (STHs). To improve this diagnostic standard, we recently designed an artificial intelligence based digital pathology system (AI-DP) for digital image capture and analysis of KK thick smears. Preliminary results of its diagnostic performance are encouraging, and a comprehensive evaluation of this technology as a cost-efficient end-to-end diagnostic to inform STH control programs against the target product profiles (TPP) of the World Health Organisation (WHO) is the next step for validation.

**Methods:** Here, we describe the study protocol for a comprehensive evaluation of the AI-DP based on its (i) diagnostic performance, (ii) repeatability/reproducibility, (iii) time-to-result, (iv) cost-efficiency to inform large-scale deworming programs, and (v) usability in both laboratory and field settings. For each of these five attributes, we designed separate experiments with sufficient power to verify the non-inferiority of the AI-DP (KK2.0) over the manual screening of the KK stool thick smears (KK1.0). These experiments will be conducted in two STH endemic countries with national deworming programs (Ethiopia and Uganda), focussing on school-age children only.

**Discussion:** This comprehensive study will provide the necessary data to make an evidence-based decision on whether the technology is indeed performant and a cost-efficient end-to-end diagnostic to inform large-scale deworming programs against STHs. Following the protocolized collection of high-quality data we will seek approval by WHO. Through the dissemination of our methodology and statistics, we hope to support additional developments in AI-DP technologies for other neglected tropical diseases in resource-limited settings.

**Trial registration:** The trial was registered on Clinicaltrials.gov (ID: NCT06055530).

**Author summary:** Millions of deworming tablets are annually administered to children to reduce the morbidity caused by intestinal worms. To monitor the progress of these large-scale deworming programs, periodic assessments are made regarding the occurrence and prevalence of intestinal worm infections. Manual examination of a stool smear through a compound microscope remains the current diagnostic standard. We recently developed a device that utilizes artificial intelligence (AI) to scan smears and recognize eggs of intestinal worms. Encouraging preliminary results of the diagnostic performance warrant additional and more research, essential for obtaining necessary approvals to support wide-scale adoption.

Here, we describe the study protocols we will employ for a comprehensive evaluation of this AI-based device. The generated results will provide health decision-makers with evidence-based data to assess whether the tool can be recommended for informing large-scale deworming programs against intestinal worms. Additionally, we provide full access to our study documentation which may be relevant for evaluating other AI-based devices for intestinal worms.

## Introduction

Soil-transmitted helminths (STHs) are a group of intestinal roundworms transmitted through the uptake of infectious life stages in the environment (often soil, referring to their common name) [1, 2]. STHs, including the giant round worm (*Ascaris lumbricoides*), whipworm (*Trichuris trichiura*) and hookworms (*Necator americanus* and *Ancylostoma duodenale*), primarily affect impoverished communities in (sub)tropical countries [1–3]. It was estimated that 24% of the global population is affected by at least one of these STHs, resulting in a total loss of 1.9 million disability-adjusted life years in 2019 [4, 5]. In response to this public health issue, many STH-endemic countries have implemented national school-based deworming programs, providing periodic oral anthelminthic treatment to the children at the schools in the program [6–8]. The pharmaceutical industry’s contribution of more than 6.5 billion anthelmintic tablets for at-risk populations since 2016 has undoubtedly contributed to reducing the disease burden in various STH-endemic countries [9, 10].

Encouraged by this progress, World Health Organization (WHO) has published its roadmap for STHs for the next decade (2020 – 2030), encompassing six ambitious targets (**Table 1**) [7, 11]. To advance towards the first two targets, it will be critical to periodically assess the STH infection prevalence, of both any intensity and moderate-to-heavy intensity (MHI) infections. The prevalence of any intensity STH infection is deployed as a parameter to determine the frequency of deworming (**Target #2**), while the elimination as a public health problem is defined when prevalence of MHI infections is less than 2% (**Target #1**) [7].

**Table 1.** The six 2030 targets and corresponding milestones put forward by the WHO [7].

| Target | Milestone |
| --- | --- |
| #1 Achieve and maintain elimination of STH morbidity in pre-school-aged and school-aged children | 98 countries with <2% children with MHI infections |
| #2 Reduce the number of tablets needed for large-scale deworming programs for STHs | 50% reduction |
| #3 Increase domestic financial support to deworm STHs | 25 countries deworming children by domestic funds |
| #4 Establish an efficient STH control program in adolescent, pregnant and lactating women of reproductive age | Coverage equals 75% |
| #5 Establish an efficient strongyloidiasis control program in school-aged children | 75% of the children at risk of *Strongyloides* receiving ivermectin |
| #6 Ensure universal access to at least basic sanitation and hygiene by 2030 in STH-endemic areas | Reduce open defecation to 0% |

Microscopic examination of a stool smear using the Kato-Katz (KK) thick smear technique and manual counting of STH eggs remain the recommended diagnostic standard for epidemiological surveys designed to inform large-scale deworming programs [7, 12, 13]. While KK thick smear is the sole diagnostic method mentioned in the 2030 targets for STHs [7], this diagnostic tool has some significant pitfalls: test results are prone to human error; it lacks clinical sensitivity when the intensity of infections is low, and hookworm eggs disappear when smears are not examined within 1h following preparation of the smear [14–17]. Within the last two decades, a variety of alternative diagnostic tools have been developed or repurposed, and subsequently evaluated for the diagnosis of STH infections in children [13, 18–21]. Despite improved clinical sensitivity for some diagnostic tools [15, 16], their integration into national deworming programs has been challenging due to labour-intensive procedures and resource demands [22]. Furthermore, as programs progress toward STH control and elimination, clinical specificity becomes increasingly more important [23]. Indeed, in the WHO’s target product profiles (TPPs) for new diagnostic tools to monitor large-scale deworming programs against STHs, the clinical sensitivity can drop to 60%, while the clinical specificity should be at least 94% [24]. The high clinical specificity of KK thick smear (≥95) [16, 25, 26] remains a strong advantage, reinforcing its likely role as a reference diagnostic for the next decade. While KK thick smear is likely to remain crucial, ongoing research and innovations in diagnostic technology show promise to address its limitations and contribute to more effective STH monitoring and control strategies [27, 28].

A clear opportunity lies in the automation of the egg counting, the step which is most prone to human error, laborious and time-demanding (egg counting takes 80% of the time-to-result, including data entry) [22]. We prototyped a proof-of-concept artificial intelligence-based digital pathology (AI-DP) device and demonstrated it for automated scanning and detection of STH eggs in KK thick smears [27]. Today, this AI-DP offers (i) electronic data capturing (EDC), (ii) whole slide imaging (WSI), (iii) an AI model and according AI development pipeline, (iv) AI results verification, and (v) a cloud-based reporting and monitoring dashboard that can be integrated into existing health systems (see also **Fig 1**). With encouraging preliminary results and field testing, a comprehensive prospective, in-the-field evaluation of the AI-DP is urgently needed to provide the necessary data for health decision makers to make an evidence-based decision on whether this technology can be recommended to inform large-scale deworming programs against STHs.

**Fig 1.**
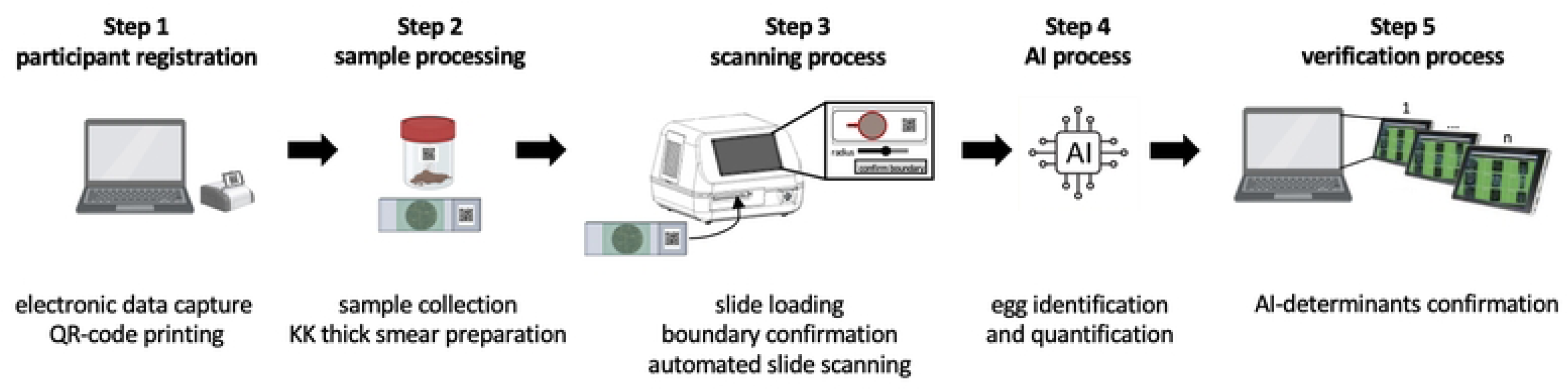
An overview of how Kato-Katz (KK) thick smears are processed with the AI-DP (KK2.0). AI: artificial intelligence, KK: Kato-Katz. Figure created using BioRender.com.

Here, we describe the study protocol for a comprehensive evaluation of an AI-DP based on its (i) diagnostic performance, (ii) repeatability/reproducibility, (iii) time-to-result, (iv) cost-efficiency to inform large-scale deworming programs, and (v) usability both in a laboratory and field setting. For each of these five attributes, separate experiments were designed to test the hypothesis that the AI-DP (KK2.0) is non-inferior when compared to the manual screening of the KK smears (KK1.0). The field work will be conducted in two STH endemic countries with a national deworming program (Ethiopia and Uganda), focussing on school-age children (SAC) only. Through the dissemination of our methodology and statistics, we also hope to support additional developments in any AI-DP technologies for other neglected tropical diseases in resource-limited settings.

## Methods

## 1 Ethics statement

The study protocol will be submitted to the institutional review boards of the Faculty of Medicine and Health Sciences of Ghent University (Belgium), the Health Institute of Jimma University (Ethiopia), the Vector Control Division Research Ethics committee (Uganda), and the Uganda National Council of Science and Technology for both review and approval. Parent(s)/guardian(s) of the participants will sign an informed consent document indicating that they understand both the purpose, and the procedures required for the study, and that they are willing to have their child participate in the study. If the child is ≥6 years old, he/she will have to orally assent to participate in the study. Participants ≥ 8 years old (≥ 12 years old in Ethiopia) will only be included if they sign an assent form indicating that they understood both the purpose of the study and the procedures required for the study, and they are willing to participate in the study. Every child that tests positive on KK1.0 or whose stool sample undergoes the egg spiking procedure will receive a single oral dose of 400 mg albendazole or 500 mg mebendazole in case of STH infections, and 40 mg/kg body weight praziquantel in case of *Schistosoma mansoni* infections. If the presence of eggs other than STHs and *S. mansoni* is confirmed, children will be referred to the nearest health centre.

The use of collected data will be strictly limited to the research objectives outlined in this study, and to enhance the accuracy and reliability of the AI diagnostic tool in identifying and diagnosing STHs. The study will adhere to the highest ethical standards, ensuring participant privacy and data protection. All data will be treated with strict confidentiality, and measures will be implemented to anonymize the data to ensure participant anonymity.

## 2 Study population and study sites

The study will focus on SAC (age 5 – 14) only, since they are the major target of large-scale deworming programs against STHs [6]. We will apply the inclusion and exclusion criteria summarized in **Table 2**. These criteria have been adapted from criteria standardized and applied throughout a series of drug efficacy trials [29].

**Table 2.**
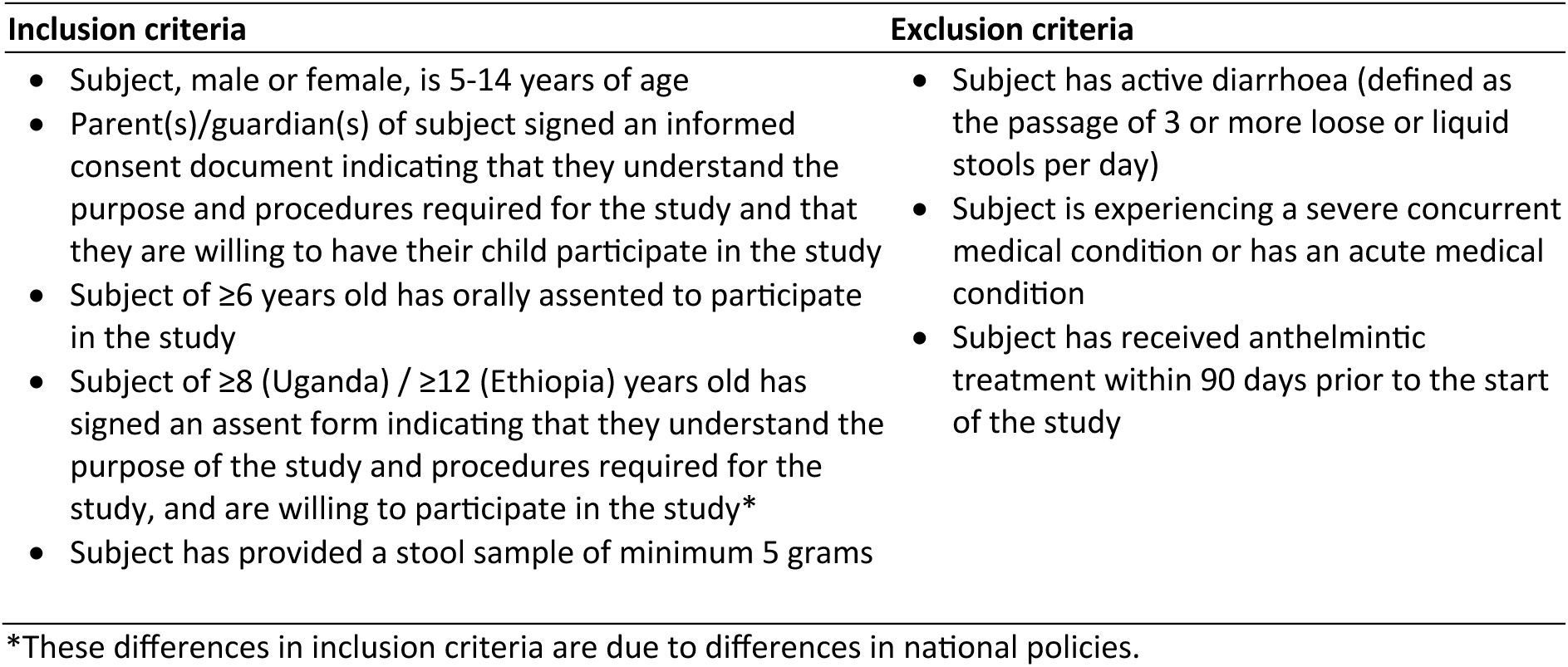
Inclusion and exclusion criteria that will be endorsed during the recruitment of participants (adapted from [29]).

The study will be conducted in both Ethiopia and Uganda. The selection of these countries and the corresponding partners (Ethiopia: Jimma University; Uganda: Vector Control and Neglected Tropical Diseases Division, Ministry of Health of Uganda) were based on ongoing collaborations [20, 29–36], the presence of an STH control program (Ethiopia: since 2015; Uganda: since 2003), and the availability of recent data on both the prevalence and intensity of STH infections [31, 32, 37]. Finally, both countries operate differently, allowing AI-DP evaluation in a fully equipped laboratory (Jimma University, Ethiopia) and a field setting (VCD, Uganda) that best mimic monitoring and evaluation (M&E) activities as part of the national STH deworming program. In Ethiopia, the study will be conducted in Jimma Zone, Oromia Regional state. In Uganda, the study will be conducted in the district of Central Uganda. The schools will be selected based on previously available data, to ensure sufficient STH cases.

## 3 Processing KK thick smears with our AI-DP (KK2.0)

Processing KK thick smears with the AI-DP (KK2.0) is graphically illustrated in **Fig 1**. To facilitate study management, the AI-DP enables EDC for registering study participants (**step 1**) and provides QR printing spreadsheets and QR label templates. Once the KK thick smears are prepared (with QR code on the slide) (**step 2**), the scanning process is initiated (**step 3**). This involves manually loading of the smears into the scanner using a specialized slide holder, after which the QR code is read, and boundary of the stool smear is determined. If required, the user is prompted to manually adjust the scan boundary. In a next step, the slide is automatically scanned, and the scanner captures focus stacks, saving eight images at every field-of-view (FOV) within the KK thick smear (**step 3**). Following slide scanning, images are transferred to the Slide Manager, and FOVs are analyzed by the AI model for the detection, classification, and quantification of helminth eggs (**step 4**). In a final step, the results generated by the AI undergo review and verification (**step 5**). This is done through the EggInspector tool, presenting all the AI-determinants from a slide to a trained verifier.

## 4 The experiments to comprehensively evaluate KK2.0

This comprehensive evaluation consists of five experiments, each one designed to evaluate one of the five attributes: (i) diagnostic performance, (ii) repeatability/reproducibility, (iii) time-to-result, (iv) cost-efficiency to inform large-scale deworming programs, and (v) usability in both a laboratory and field setting. **Table 3** provides an overview of the hypotheses, the primary and secondary outcomes for each experiment separately. Across these five experiments, we defined 9 hypotheses, 13 primary and 17 secondary outcomes. Generally, we hypothesize that KK2.0 is non-inferior to KK1.0. Note that a hypothesis was not defined for both the time-to-result and usability experiments. This was because the outcomes of the time-to-result experiment will feed into the experiment on cost-efficiency and because the usability experiment was designed to gain insights into how we can further improve the usability of KK2.0 only. In the following sections we will discuss each experiment in detail. The sample size calculation and the statistical data analysis will be discussed in **sections 2.5.** and **2.6**, respectively.

**Table 3.**
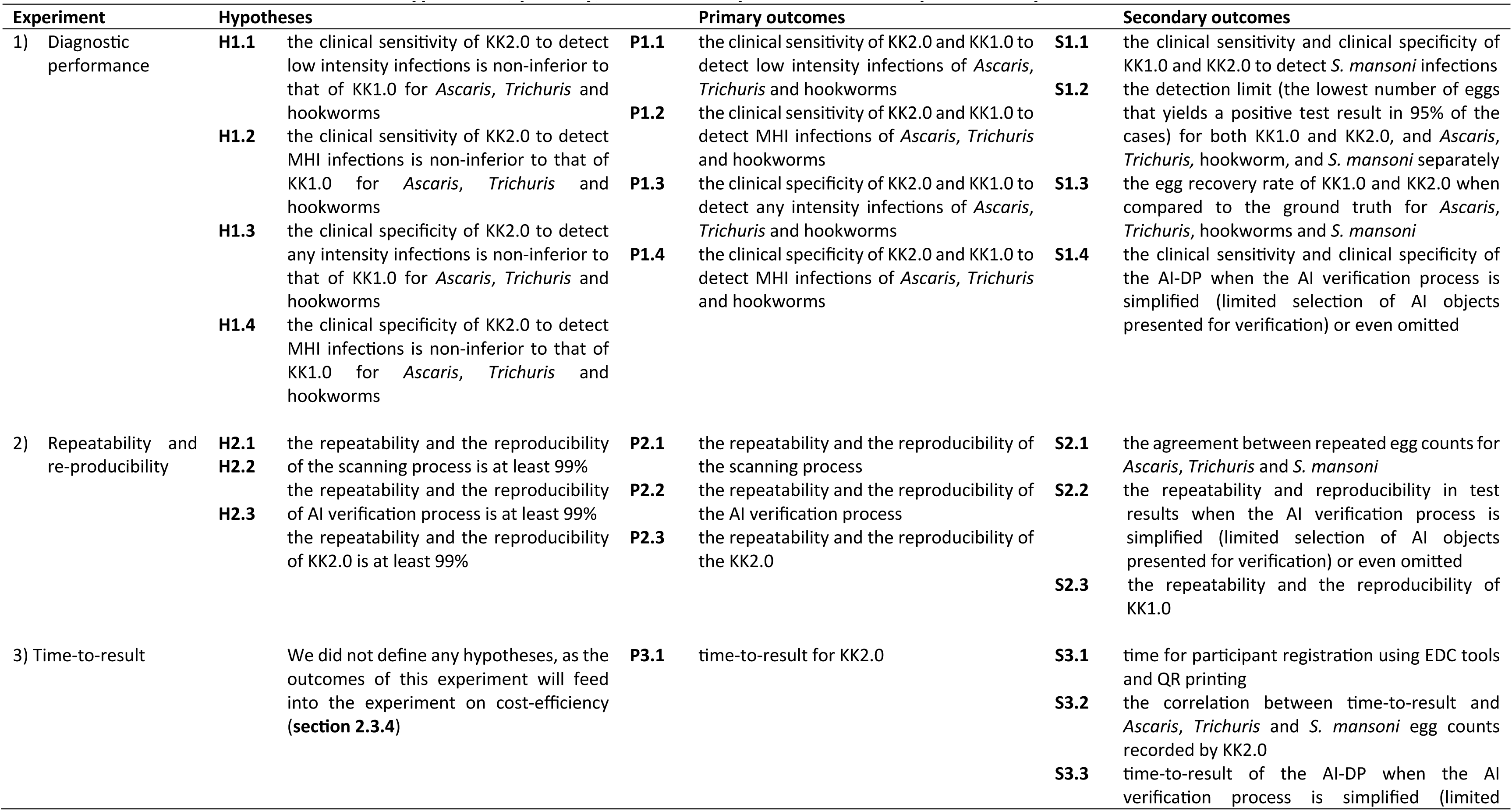
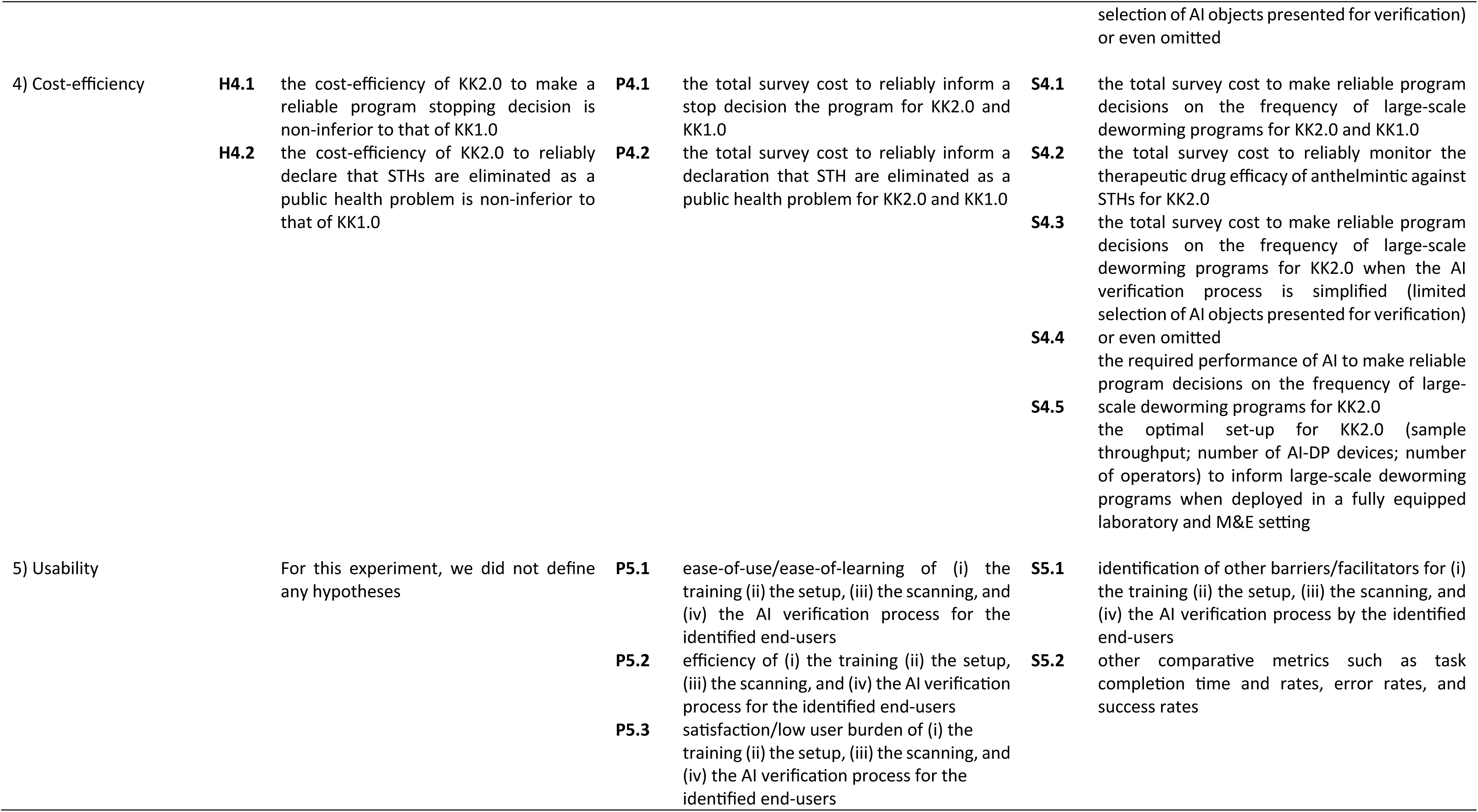
An overview of the hypotheses, primary, and secondary outcomes to comprehensively evaluate KK2.0.

### 4.1 Diagnostic performance

**Fig 2** provides an overview of the proposed study design for the experiment on the diagnostic performance. Generally, this experiment consists of five consecutive steps, with the second step offering two methods to validate diagnostic performance. The first method involves verifying egg counts by reviewing and counting all eggs within the captured FOVs. The second method entails spiking a minimum number of eggs into randomly selected stool samples to achieve counts indicating an MHI infection. In the **first step** of the experiment, fresh stool samples will be collected from SAC at the schools. In the **second step**, the consistency of the stool samples will be scored based on the Bristol Stool Chart [38]. Subsequently, sample will be homogenised, and one KK thick smear per sample will be prepared in one of the two following ways for the two validation methods described above. For FOV-based validation (**step 2A**), samples will be processed as recommended by WHO. For egg spiking-based validation (**step 2B**), the cone of stool (after removing the KK template) will be spiked with purified eggs to artificially increase the egg counts to at least an MHI infection (*Ascaris*: >209 eggs; *Trichuris*: >42 eggs; hookworms: >84 eggs). This step 2B will only be done in a subset of the samples (never on samples that are processed through 2A) and is introduced to ensure that sufficient cases of MHI infections for each of the STHs are obtained (see also **section 2.5**). The selection of the samples to be spiked will be done through a randomization process. In the **third step** and following a smear clearing time of 30 min, the smears will be randomly allocated to be analysed by either KK1.0 (even participant ID) or KK2.0 (odd participant IDs). This randomization process is required to avoid systematic bias due to hookworm egg degradation over time [13, 14, 39]. In the **fourth step**, egg counts will be recorded for each helminth species (*Ascaris*, *Trichuris*, hookworm and *S. mansoni*), separately. Thereafter (**step 5**), KK thick smears will be stored at 4°C to be used in the context of the experiment on reproducibility/repeatability (see **section 2.4.2**). In Ethiopia, sample processing (from step 2 onwards) will be conducted in the Neglected Tropical Disease Laboratory of Jimma University (a fully equipped laboratory setting), while in Uganda all steps will be conducted on-site (M&E setting).

**Fig 2.**
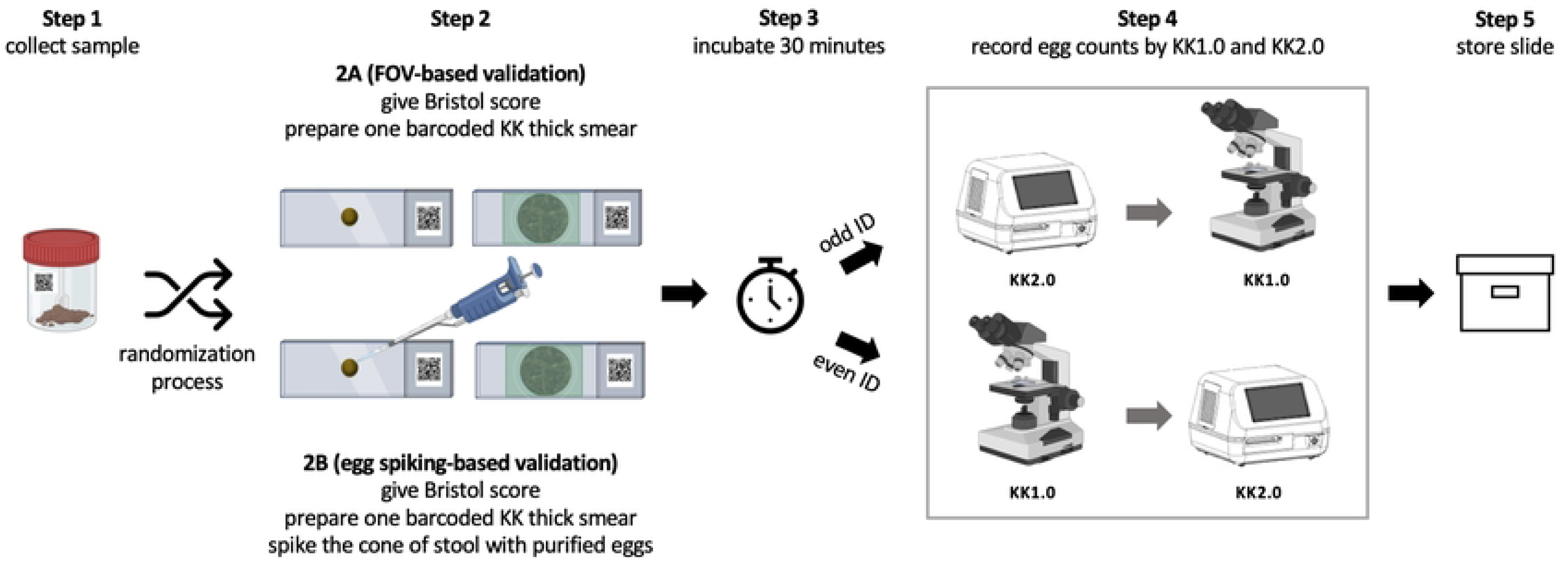
Overview of the study design for the experiment on diagnostic performance. FOV: field-of-view, KK: Kato-Katz. Figure created using BioRender.com.

In absence of a gold standard, it will be important to define the ground truth for each slide separately, to test the hypotheses (H1.1 – H.1.4). For the slides that were not spiked, all FOVs that were captured through the AI-DP will be manually annotated by one trained laboratory technician. A second trained laboratory technician will then verify the annotations. In case of disagreement, a third trained laboratory technician will make the final call. For the spiked samples, the ground truth (samples being classified as MHI infection) is already established through the process of spiking.

### 4.2 Repeatability and reproducibility

In this experiment, we will be evaluating the two parameters repeatability and reproducibility. Repeatability refers to the variability in test results (*Ascaris*, *Trichuris* and *S. mansoni*) when the same KK thick smear is examined by the same operator (e.g. scanner of AI-DP/microscopist), so called intra-annotator agreement, while reproducibility refers to the variability in test results when the same slide is examined by a different operator (e.g. scanner of AI-DP/microscopist), so called inter-annotator agreement (see also **Fig 3** for graphic definition of both repeatability and reproducibility). For KK2.0, we will focus on the scanning process (step 3 in **Fig 1**) and the AI verification process (step 5 in **Fig 1**). For KK1.0, we will focus on the egg counting process only (see also **Fig 3**). Generally, we hypothesise that both the repeatability and reproducibility of KK2.0 is at least 99%.

**Fig 3.**
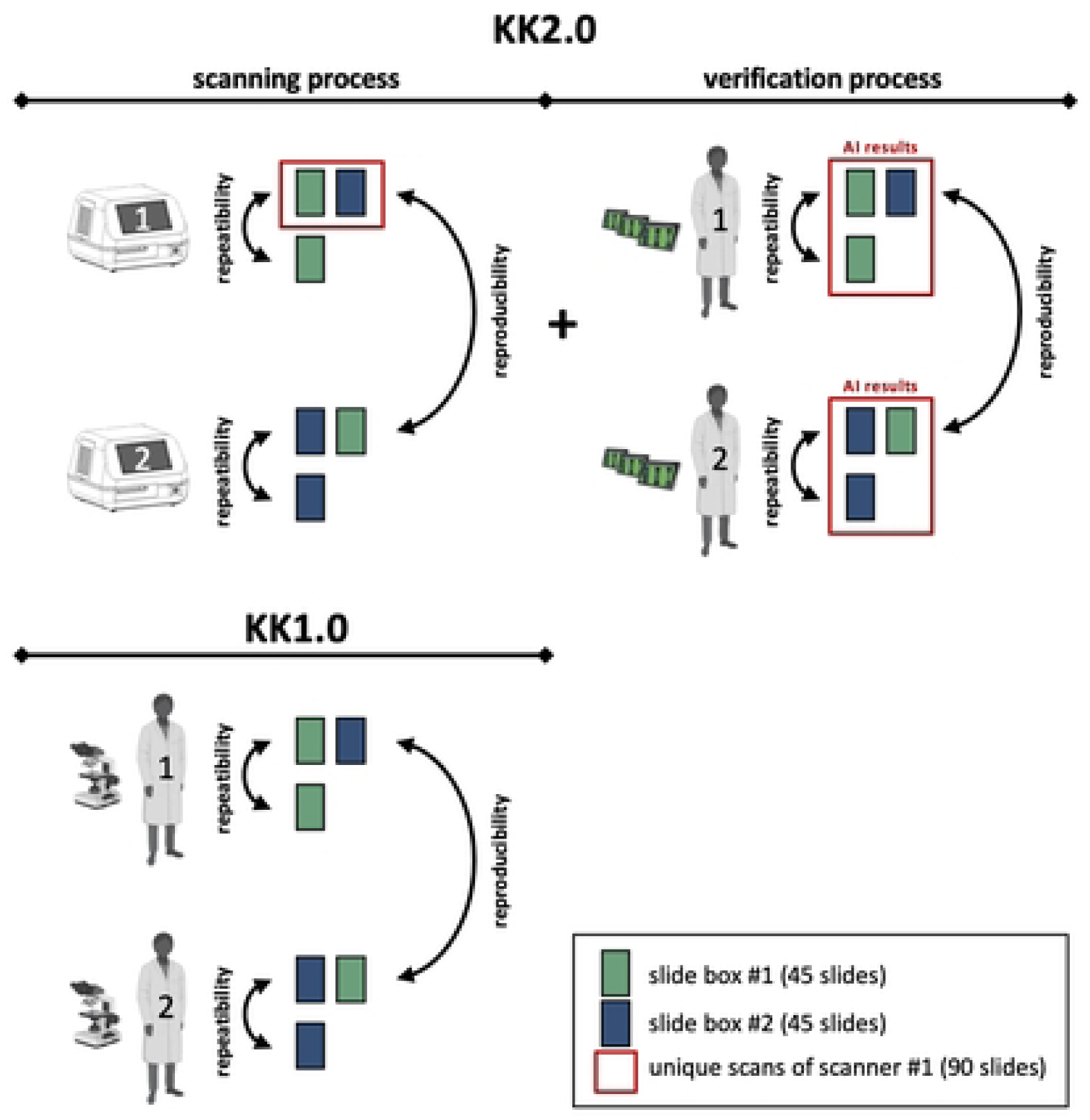
Overview of the study design for the experiment on the repeatability and reproducibility. Figure created using BioRender.com.

**Fig 3** provides an overview of the proposed study design for the experiment on the repeatability and reproducibility. For this experiment, we will use a subset of the KK thick smears prepared during the experiment on the diagnostic performance. The subset will comprise two slide boxes, each containing 45 KK thick smears. To ensure we assess the repeatability and reproducibility across different egg counts, we will randomly select 30 negative KK thick smears, 30 smears with a total egg count for any helminths (*Ascaris*, *Trichuri*s and *S. mansoni*) between 1 and 100, and 30 smears with a total egg count greater than 100, resulting in a total of 90 smears. We will ensure that at least 50% of the KK thick smears in each box contain eggs from at least two different helminth species.

For the repeatability and reproducibility of the scanning process (KK2.0), all KK thick smears in slide box #1 (green) and box #2 (blue) will undergo two rounds of scanning. To ensure the entire sample is scanned, boundaries will be set larger than the smear for every scan, limiting interference and error caused by human error. The repeatability and reproducibility of the scanner process will be based on the final test results generated by the complete scanning process, which includes slide loading, boundary setting, device calibration, automatic focus setting, scan algorithms, AI detections and egg grouping algorithms. For the repeatability and reproducibility of the result verification process, the AI results of the unique scans of scanner #1 (red frame) will be verified by at least two different microscopists. For KK1.0, the examination of the KK thick smears will be conducted using the same flow as applied for the AI verification process. We will ensure that the same microscopists examine the same slides for both KK1.0 and do the AI verification for KK2.0.

### 4.3 Time-to-result

During the experiments on the diagnostic performance (**section 2.4.1**), and repeatability and reproducibility (**section 2.4.2**), four different steps of the KK2.0 procedure will be timed. The four steps involve (i) participant registration (step 1 in **Fig 1**), (ii) the scanning process (step 3 in **Fig 1**), (iii) the AI process (step 4 in **Fig 1**), and (iv) the verification process (step 5 in **Fig 1**). The time required for each of these steps to be completed will be recorded by the AI-DP. The total time-to-result will be defined as the sum of the durations needed for the individual steps. Furthermore, in the Ugandan field setting, the time for setting up the AI-DP system at the different field locations will be recorded. The time-to-result for KK1.0 will not be measured in the present study. This has been intensively researched elsewhere as part of four clinical trials, each trial conducted in a different country [29, 40]. We will use these data as a comparator for KK2.0.

### 4.4 Cost-efficiency

For this experiment, we built up on two general frameworks that were previously developed to support cost-efficient study design choices for large-scale STH deworming programs, including epidemiological surveys to reduce/stop large-scale deworming programs and to declare STH eliminated as a public health problem [41, 42], and to monitor the therapeutic drug efficacy [22]. Generally, these frameworks consist of three consecutive steps. In the **first step**, an in-depth analysis of the operational costs to process one stool sample is conducted for each diagnostic tool. In the **second step**, simulation studies are performed to determine the probability of making the reliable program decision. In the **third step**, the outcome of the cost assessment is integrated into the simulation study to estimate the total survey costs and determined the most cost-efficient study design. For the in-depth analysis of the operational costs to process one sample, we will both conduct an itemized cost assessment and determine the salary costs, which will be a function of the time-to-result (see **section 2.4.3**). For the simulation, we will deploy simulation frameworks previously published by both Kazienga et al. (2023) and Coffeng et al. (2023) [22, 41]. Both frameworks account for different sources of variation in egg counts, including (i) variability in mean egg intensity between schools; (ii) inter-individual variability in mean egg intensity due to variation in infection levels between individuals, where the level of aggregation is a linear function of the school-level mean egg intensity; (iii) day-to-day variability in mean egg intensity within an individual due to heterogeneous egg excretion over time; (iv) variability in egg counts between repeated aliquots of a stool sample due to the aggregated distribution of eggs in stool; (v) inter-individual variability in the effect of drug administration. Through the outputs of the experiment on both the diagnostic performance (**section 2.4.1**), and the repeatability and reproducibility (**section 2.4.2**), we will be able to further customize the simulation work to KK2.0 (e.g., additional variation in test results due to AI verification process and imperfect egg recovery).

### 4.5 Usability

We define usability as the degree to which the KK2.0 can be used easily, efficiently, and with satisfaction/low user burden by the stakeholders [43]. For this experiment, KK2.0 naïve participants (having no previous exposure or experience with the system) will receive practical training in the use of the KK2.0 system, which includes three steps, namely the set-up, the scanning, and the AI verification process.

The practical training consists of an initial demonstration of this three-step process and a walk-through of system user manuals. Afterwards, the participants will be invited to two natural use environments, either to a laboratory setting, or a field setting. The participants will be organized into four groups per setting, each consisting of two participants per group, resulting in a total of 16 participants. This grouping reflects a planned real-life group setup, wherein the involvement of two laboratory technicians are expected to carry out the tasks. The group will be asked to perform the set-up as a team. The two following steps, the scanning and verifying AI, will be performed individually. For this, participants will be asked to each process 6 slides with KK2.0. Each slide will be processed in following order, whereby the participant’s effort will be increased: (1) the final results are available soon after scanning is complete (e.g., KK2.2 results), (2) the user must perform the simple verification procedure before the results are available, (3) the user must perform the complete verification procedure before the results are available. During the three-step task performance, participants will verbalize their experiences and detect weak points in their interaction with the scanner (i.e., think–aloud protocol [44]). The whole session will be video-recorded, and data will be generated by verbatim transcriptions, and an observation checklist for collecting comparative metrics (e.g., task completion time and both error and success rates). Following, a semi-structured interview will be implemented to capture the ease-of-use/ease-of-learning, efficiency, and satisfaction/low user burden, as well as potentially missed barriers and facilitators during the task completion process. The interviews will be conducted by one investigator and structured around four sections: the background of the participant; the training; the KK2.0; the context. The data will be audio-recorded and transcribed verbatim.

## 5 Sample size calculation

A formal sample size calculation was conducted for the experiments on the diagnostic performance (**section 2.4.1**), and the repeatability and reproducibility (**section 2.4.2**). For the other experiments we did not determine the sample size, because either no hypothesis was defined as the outcomes will feed into another experiment (**section 2.4.3 Time-to-result**), the hypothesis is based on a simulation study (**section 2.4.4 Cost efficiency**), or the sample size was based on common practice in literature (**2.4.5. Usability**). In the following sections we will only briefly discuss the applied methodology to determine the sample size for the three experiments (diagnostic performance, repeatability/reproducibility, and usability). For a detailed description of the applied methodology for the first two experiments we refer the reader to **S1 Info.**

### 5.1 Diagnostic performance, repeatability, and reproducibility

Generally, we opted to conduct a series of simulation studies over the standard sample size methodologies, as this approach allowed us (i) to better capture the variation in test results that are otherwise difficult to account for (e.g., clinical sensitivity of KK1.0 increases as a function of egg numbers in a slide), and (ii) to ensure that the sample size calculation and the final interpretation of the field data are both based on the same statistical approach (e.g., the relative position of confidence intervals (CI) to predefined set of values; see also **Fig 4**). In brief, each of these simulation studies consists of a series of in-silico experiments that are iterated under different conditions (e.g., different sample sizes). Based on this iterative process, we determined the lowest sample size that allowed for confirming the hypothesis in at least 80% of the iterations (= power).

**Fig 4.**
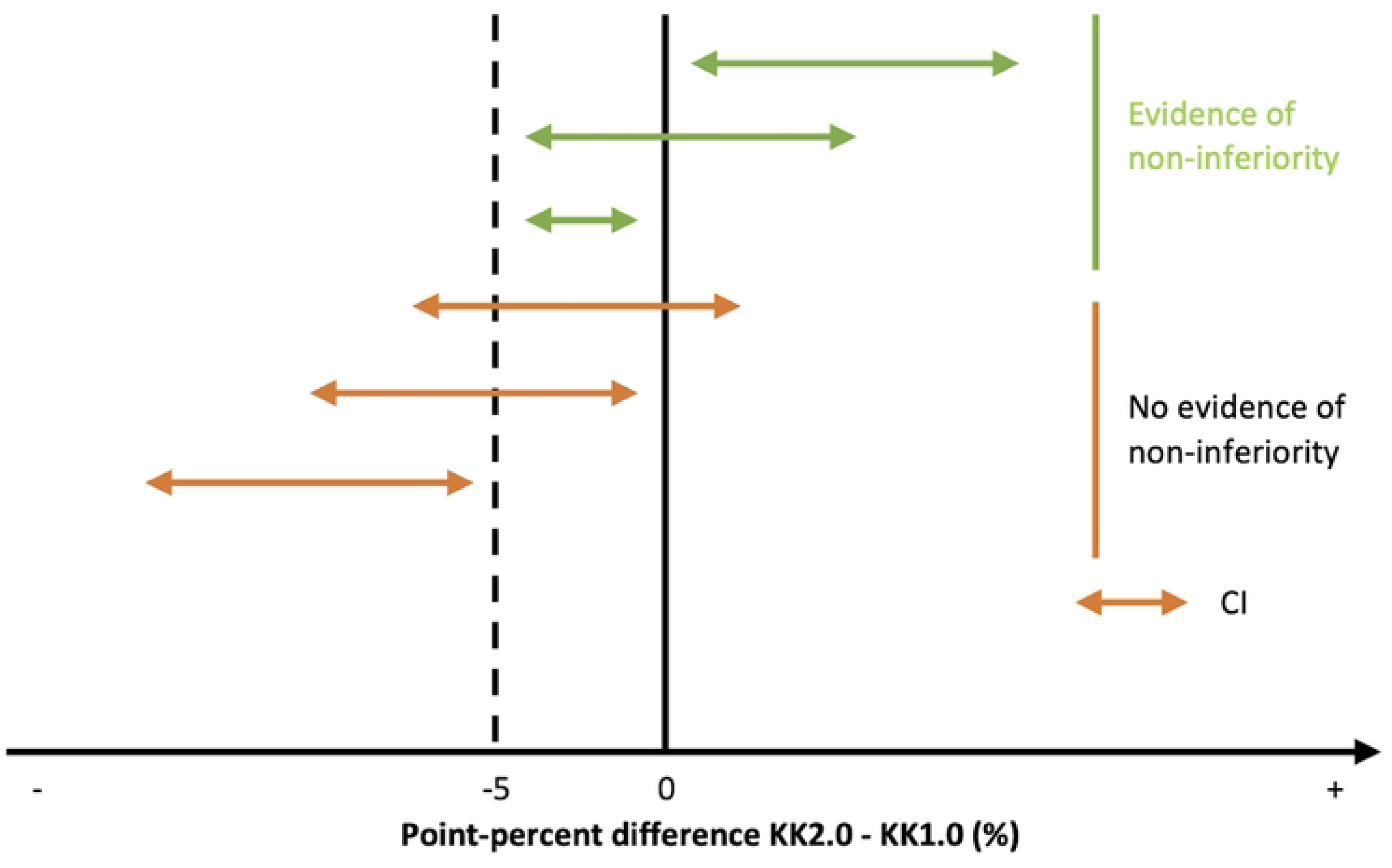
Overview of the different outcome scenarios based on a random sample and its corresponding CI. This figure illustrates the different outcome scenarios around the difference in performance between KK2.0 and KK1.0 based on the CI. The green lines represent the scenarios where there is evidence of non-inferiority, while the lines in orange illustrate the scenarios where there is no evidence of non-inferiority. In this example we set the level of equivalence at −5 percent difference between (KK2.0 – KK1.0), a negative value indicating that KK1.0 is better.

#### 5.1.1 Diagnostic performance

For the clinical sensitivity to detect low intensity (***H1.1***) and MHI infections (***H1.2***), we accounted for (i) a varying clinical sensitivity as a function of the number of eggs in a slide; (ii) a proportion of the eggs in a slide being missed, (iii) correlation between test results of KK1.0 and KK2.0 on the same slide, (iii) and helminth specific FEC thresholds defining low intensity and MHI infections (**Table 4**). In this simulation, we assumed that the clinical sensitivity of KK2.0 is equal to that of KK1.0 and an equivalence level of 5-point percent. In other words, the lower limit of the CI around the difference (KK2.0 – KK1.0) should be at least −5% (see also **Fig 4**). As we will draw conclusions on three different STHs at the same time and because we are testing for non-inferiority, we set the level of significance at 0.05/3.

**Table 4.** The FEC thresholds defining low intensity and MHI STH infections. This table summarizes the WHO FEC (in EPG) thresholds to classify the intensity of STH infections into low, moderate and heavy [45].

| Helminth | Low | Moderate | Heavy |
| --- | --- | --- | --- |
| *Ascaris* | 1 – 4,999 | 5,000 – 49,999 | ≥50,000 |
| *Trichuris* | 1 – 999 | 1,000 – 9,999 | ≥10,000 |
| Hookworm | 1 – 1,999 | 2,000 – 3,999 | ≥4,000 |

**Table 5.**
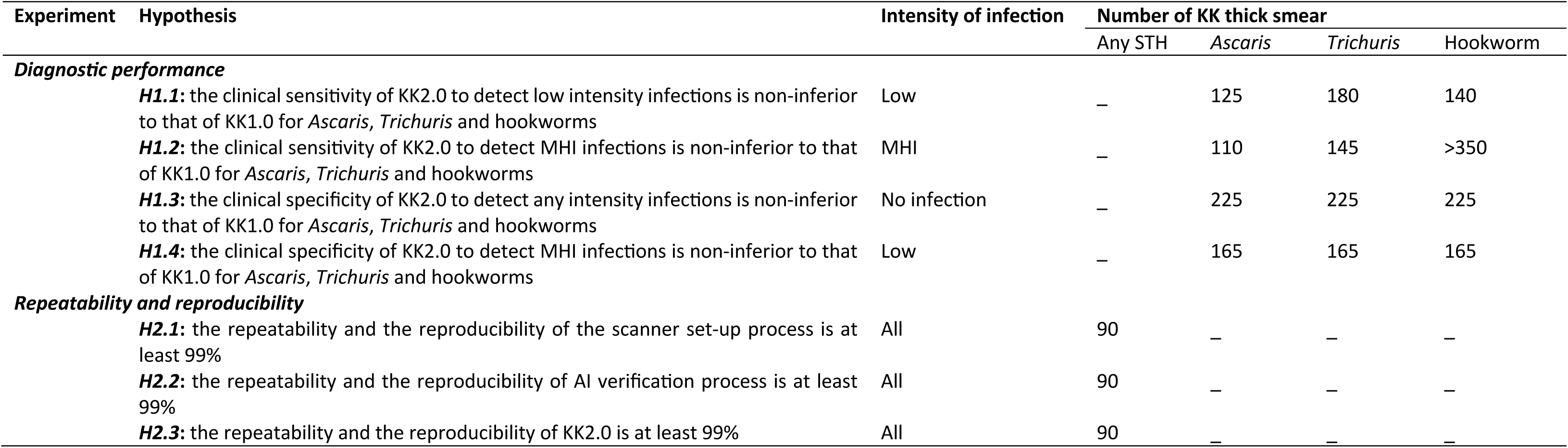
Overview of the required number of KK thick smear to test the hypotheses for the experiments on diagnostic performance and repeatability/reproducibility.

Based on these assumptions, the required number of KK thick smears representing low intensity infections based on the ground truth is 125 for *Ascaris*, 180 for *Trichuris* and 140 for hookworms. The required number of KK thick smears representing MHI infections based on the ground truth, is 110 for *Ascaris* and 145 for *Trichuris*. For hookworms, the required sample size exceeded 350, which revealed to be beyond the capacity of this project.

For the clinical specificity to detect any intensity (***H1.3***) and MHI infections (***H1.4***), we used another data generation process (based on binary test results (positive/negative) instead of egg counts). Because of this, the required sample size is the same for each of the different STHs. In this simulation study, we also (i) accounted for correlation between test results of KK1.0 and KK2.0 on the same KK thick smear, (ii) assumed an equal clinical specificity for both diagnostic tools, an equivalence level of 5-point percent, and (iii) set the level of significance at 0.05/3. Based on these assumptions, the required number of KK thick smears representing no infections based on the ground truth is 225 for *Ascaris*, *Trichuris* and hookworms each. Consequently, the required number of KK thick smears representing low intensity infections based on the ground truth is also 165 for each of the three STHs separately.

#### 5.1.2 Repeatability and reproducibility

To verify whether the repeatability and reproducibility for the scanner set-up (***H2.1***), the AI verification process (***H2.2***), and the complete KK2.0 (***H2.3***), is at least 99%, we conducted a simulation study where we determined the number of KK thick smears that resulted in a lower limit of the CI that is at least 95% in 80% (= power) of the iterations when the true underlying probability of success equals 99%. Given that we are testing both repeatability and reproducibility at same time for each process, and that we are testing for non-inferiority, we set the level of significance at 0.05/2. Based on these assumptions the, required KK thick smears that need to be re-processed equals 90 for each of the three hypotheses.

### 5.2 Usability

In this experiment, we will include 16 participants to receive (i) practical training and engage in the three-step process (ii – iv) and usability testing. A group size of 3-20 participants is considered valid in such problem discovery scenarios, with 5-10 participants being a sensible baseline range [46]. The group size should typically be increased along with the study’s complexity and the criticality of its context. Since the study will take place in two different settings, either in a well-equipped laboratory or field setting, we considered 8 participants per setting, resulting in a total of 16 participants (4 groups of 2 participants per setting).

## 6 Statistical data analysis

### 6.1 Diagnostic performance

#### 6.1.1 Primary outcomes

We will draw contingency tables representing the test results of both KK1.0 and KK2.0 for each type of ground truth (no, low intensity and MHI infections) and STH species (*Ascaris*, *Trichuris* and hookworms). From these tables, both the clinical sensitivity and specificity, and the corresponding 95% CI (Wald) will be calculated for each test and STH separately. Subsequently, we will also calculate the 90% CI around the difference in performance (KK2.0-KK1.0). Given that test results are paired (same smears are processed by KK1.0 and KK2.0, we will use the formulae described by Newcombe for paired data [47]. We will conclude that the clinical sensitivity or specificity of KK2.0 for a particular STH is non-inferior if the lower limit of the 90% CI does not include the −5-point percent.

#### 6.1.2 Secondary outcomes

We will draw contingency tables representing the test results of both KK1.0 and KK2.0 for each type of ground truth for *S. mansoni* infections. From these tables, both the clinical sensitivity and specificity, and the corresponding 95% CI will be calculated (***S1.1***).

To determine the detection limit (the lowest number of eggs that yields a positive test result in 95% of the cases) of KK1.0 and KK2.0 for STH and *S. mansoni* (**S1.2**), logistic regression models accounting for repeated measures will be built for each helminth species separately using the ‘mixed_model’ function in R. The test result (positive or negative) will be used as dependent variable while ‘test’ (2 levels: ‘KK1.0’, ‘KK2.0’), log transformed egg counts based on ground truth at first examination, Bristol stool scale and all two-way interactions will be used as predicting variables. From these models, we will predict the probability having a positive test result and the corresponding 95% prediction interval for each integer value of ground truth egg counts between 1 and 100 using the ‘marginal_coefs’ function in R. We will define the detection limit as that range of egg counts for which the 95% prediction intervals include 0.95. We will explore the egg recovery rate (= observed egg counts / ground truth egg counts) of KK1.0 and KK2.0 when compared to the ground truth for *Ascaris*, *Trichuris*, hookworms and *S. mansoni* (***S1.3***). These analyses will only be conducted on KK thick smears representing low intensity infections. Finally, we will draw contingency tables representing the test results of KK2.0 for each type of ground truth (negative, low intensity and MHI infections) for each helminth species and AI verification process (simplified AI verification (limited selection of AI objects presented for verification) *vs.* no AI verification), separately. From these tables, both the clinical sensitivity and specificity, and the corresponding 95% CI will be calculated for each helminth species and type of AI-verification process (***S1.4***).

### 6.2 Repeatability and reproducibility

#### 6.2.1 Primary outcomes

The egg counts on the same smear will be considered not repeatable/reproducible in one of the following three scenarios of discrepancy: (i) there is a difference in presence/absence, (ii) the difference in egg counts exceeds 10 eggs for slides with egg counts *≤*100 eggs, (ii) the difference in egg counts exceeds 20% eggs for slides with egg counts >100 eggs. These criteria are developed by the Swiss Tropical Institute of Tropical and Public Health (Speich et al., 2015), and are currently the standard way of quality control of egg counts in clinical trials [25, 26].

To determine the repeatability (proportion of cases for which a repeated test result by the same operator/scan met the aforementioned criteria) and reproducibility (proportion of cases for which a repeated test result by a different operator/scan met the aforementioned criteria) of the scanning process (***P2.1***) and AI-verification (***P2.2***), we will draw contingency tables representing the repeated test results of KK2.0 on the same KK thick smears by the same operator / scanner (repeatability) or different operator / scanner (reproducibility) for each of the different steps of the KK2.0. From these tables, both the repeatability and reproducibility, and the corresponding 90% CI (Wald) will be calculated for the scanning process, AI-verification and complete KK2.0, separately. We will conclude that the reproducibility/repeatability of these steps are at least 99% if the 90% CI does not include 95%.

#### 6.2.2 Secondary outcomes

We will explore the agreement in repeated egg counts by using a Bland-Altmann plot for the scanning process, AI-verification, the complete KK2.0 and KK1.0 for each of the three helminths, separately (***S2.1***). In addition, we will repeat the analysis of repeatability and reproducibility for both a simplified AI-result verification process (limited selection of AI objects presented for verification) and where AI-result verification is omitted (***S2.2***).

### 6.3 Time-to-result

We will determine the mean (and corresponding 95% confidence intervals) time-to-result (***P3.1***) and the time for participant registration using EDC tools (***S3.1***). In addition, we will also explore the correlation between time-to-result and *Ascaris*, *Trichuris* and *S. mansoni* egg counts recorded by KK2.0 (***S3.2***) based on the Spearman’s coefficient. Finally, we will repeat the analysis to determine the time-to-result of our AI-DP when the AI verification process is simplified (limited selection of AI objects presented for verification) and where AI-result verification is omitted (***S3.3.***)

### 6.4 Cost-efficiency

We refer the reader to **section** 2.4.4.2 for more details.

### 6.5 Usability

To achieve a thorough comprehension of the training and scanner usability, we will employ data triangulation as a method for analysing and incorporating multiple data sources. The approach to qualitative data analysis will combine inductive and deductive elements, using the determinants of usability: ease-of-use; efficiency; satisfaction/low user burden. Analytical categories will be developed from the initial research questions and emerge during the analysis process. Using NVivo (Version 14, 2020, Lumivero), identified categories will be operationalized as codes in a flexible coding scheme. The content of the codes will be discussed extensively between independent coders, and subsequently used to identify pain points and to explore improvements. The quantitative data obtained through the observational checklists will be analyzed through basic descriptive statistics.

## Discussion

Despite the well-known limitations of KK thick smear, it is probably here to stay for the next decade. As response to this, we have designed and developed an AI-DP (KK2.0) that could overcome some of these limitations. Moreover, by incorporating both EDC tools and cloud-based reporting with a monitoring dashboard that can be integrated into existing health systems, KK2.0 holds promise as an end-to-end diagnostic tool in large-scale deworming programs targeting STH. Encouraged by preliminary results on the diagnostic performance, we now want to provide the data necessary to make more evidence-based decisions on the potential of this AI-DP.

## 1 Comprehensive evaluation beyond diagnostic performance

While the evaluation of new diagnostic methods has often been limited to the clinical sensitivity and specificity only, we deliberately opted to evaluate additional attributes and combine them into a simulation study that is designed to determine the cost-efficiency of the AI-DP to inform large-scale deworming programs. As recently illustrated for monitoring the therapeutic efficacy against STHs [22], we strongly believe that this holistic approach is required to make any evidence and value-based decisions. This is particularly relevant for STH control programs which operate in resource poor settings, and hence it will be important to ensure reliable and confident programmatic decision making, while minimizing the operational costs. Moreover, a complex interplay exists between the diagnostic performance and the epidemiological setting (e.g., clinical sensitivity reduces in low endemic setting [15, 41], the sample throughput, and the operational costs (e.g., improving the diagnostic performance and the corresponding reduced sample sizes can compensate for more costly tests and lower sample throughput; there is a limit to the extent to which higher reagent costs can be compensated by lower sample throughput) [23, 42]. In other words, it would be quite impossible to draw conclusions on whether any new diagnostic method holds promise to inform large-scale deworming programs without fully exploring these aspects in more detail [22, 41]. On top of these, we have set-up a usability experiment, to further adjust the AI-DP to user’s requirements.

## 2 Estimates of diagnostic performance are not absolute, but relative to KK1.0

For many infectious diseases, the absence of a gold standard (100% sensitivity and specificity) is a universal challenge to estimate the true performance of new diagnostics [48, 49]. To overcome this obstacle for STHs, it has been suggested to examine more stool samples with multiple diagnostic methods [50–53], and to deploy statistical methodologies that account for the absence of a gold standard [49]. In our study, we will determine the diagnostic performance of the AI-DP relative to the current diagnostic standard (KK1.0). In our opinion choosing KK1.0 as a sole comparator is justified. First, the AI-DP aims to improve the current KK1.0, and hence it is the obvious comparator to test the non-inferiority hypotheses. Second, for MHI infections, KK1.0 remains the sole diagnostic method to define the intensity of infections [23, 24]. Third, it has recently been shown that the clinical specificity, rather than the clinical sensitivity, will become more important when programs progress towards control and elimination of STH [23, 54]. Clinical specificity of KK1.0 thick smear (95% [16, 25, 26]) has never been considered as a drawback, which takes away the need for a more sensitive comparator (e.g., qPCR [15, 55]). Finally, we carefully designed the experiments so that we can ensure the true underlying infection status. For the KK smears representing no infections or infections of low intensity, we will have the ground truth based on the scans of the KK thick smears, while for the smears representing MHI infections we will spike the slides with known number of eggs. This design allows us to draw the appropriate conclusions around the defined non-inferiority hypotheses without the need of other diagnostic methods (e.g., qPCR) or more complex statistical models that account for a gold standard.

## 3 Alignment with WHO TPPs for STHs

In 2021, WHO published its TPP for STH, defining the minimal and ideal criteria for 38 attributes organized in five clusters (product use summary: 5 attributes; design: 11 attributes; performance: 10 attributes; product configuration: 5 attributes; product cost and channels: 5 attributes)[24]. A year later, we systematically analysed this TPP for an AI-DP solution [27]. **Fig 5** provides a graphical overview per cluster of how the current AI-DP already meets these criteria, and for which attributes this study will provide full, partial or no evidence. In S2 Info, we provide the same information for each attribute separately. Today, our AI-DP already meets 14 attributes and through this study we will provide partial or full evidence for another 17 attributes. The study will not address the remaining 7 attributes because they are considered to be out of scope. Most of these attributes are within product configuration (shipping conditions and labelling and instructions for use), and product cost and channels (product lead times, target launch countries and product registration), and therefore will need to be addressed at a later stage when there is sufficient evidence that our AI-DP meets the other attributes. Note that, the reproducibility and repeatability is not considered as an attribute in the WHO TPP.

**Fig 5.**
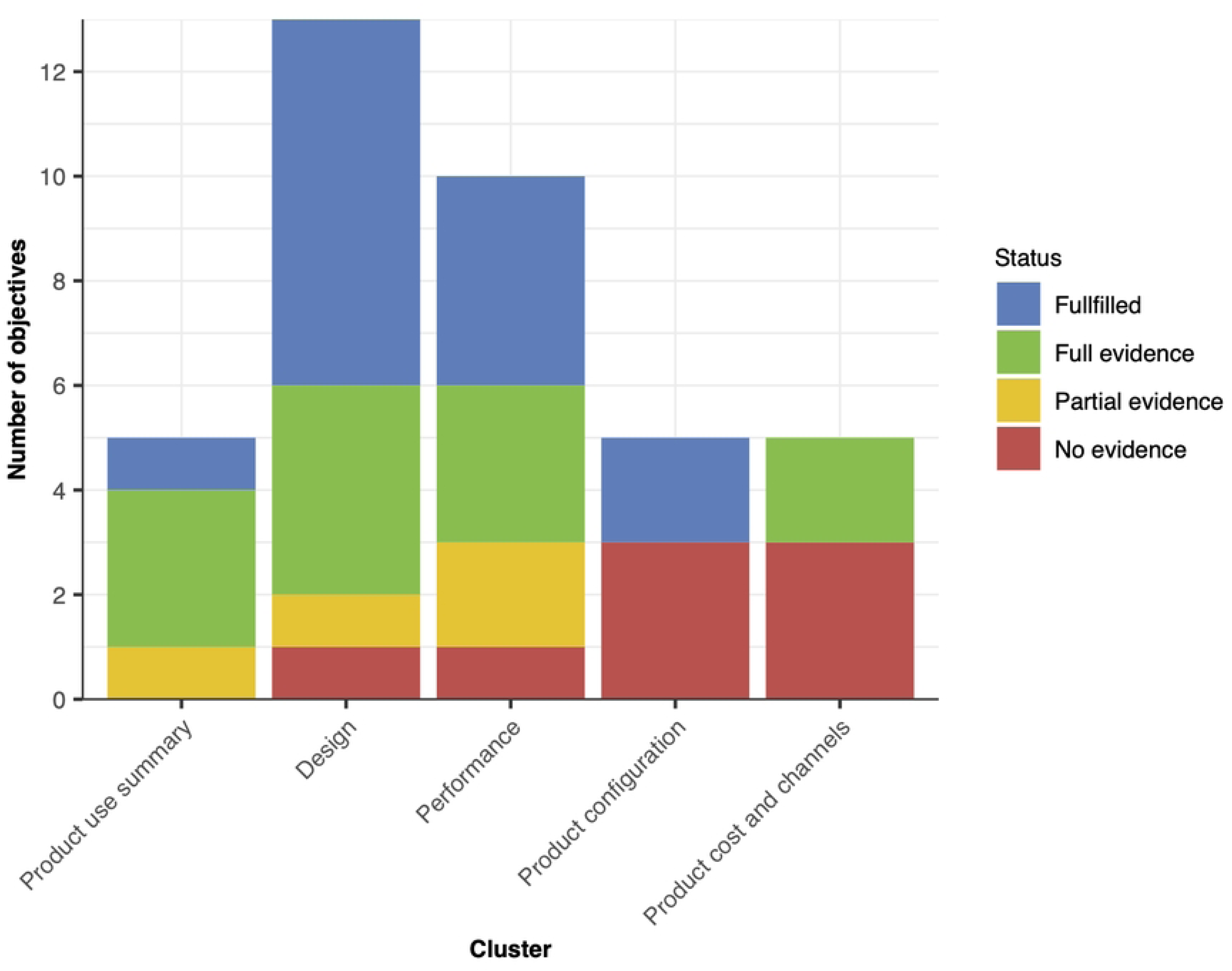
Overview per cluster of how the current AI-DP already meets the attributes defined in the WHO TPP criteria, and for which attributes this study will provide full, partial or no evidence.

## 4 Moving from KK2.0 over KK2.1 to K2.2

Today, the AI-DP still relies on the human operator to verify all the detections by AI (KK2.0). It is our ambition to further minimize this in two consecutive steps. In first step, we will reduce the number of detections presented for human verification, e.g., to the detections for which there is doubt (KK2.1). In a final step, all human verification will be removed, and results will rely on AI only (KK2.2). During this study, we will already gather the evidence for both KK2.1 and KK2.2 (secondary outcomes *S1.5*, *S2.2*, *S3.3*, *S4.3*; see Table 3). Moreover, through the usability experiment we will be able to further customize the AI-DP and corresponding needs of the key end-users.

## Conclusions

This comprehensive study will provide the necessary data to make an evidence-based decision on whether our AI-DP is indeed a cost-efficient end-to-end diagnostic to inform large-scale deworming programs against STHs. In case of a favourable outcome, we will seek further guidance by WHO. Meanwhile, we provide full access to sample size calculations and record forms, which may be relevant for the evaluation of any other AI-DP or diagnostic.

## Data Availability

All relevant data are within the manuscript and its Supporting Information files.

## Supplementary info

**S1 Info. The methodology to determine the required sample sizes to test the project hypotheses around diagnostic performance, repeatability, and reproducibility.**

**S2. Info. Detailed overview of how our current AI-DP already meets the attributes defined in the WHO TPP criteria, and for which attributes this study will provide full, partial or no evidence.**

## Acknowledgements

We are extremely grateful to Dr. Lieven Stuyver (Janssen Global Public Health, Janssen R&D, 2340 Beerse, Belgium) for initiating the concept of an AI-DP for NTDS, and to continue steering multi-disciplinary teams worldwide towards a proof-principle for the AI-DP described in this work.

